# Outcome Risk Modeling for Disability-Free Longevity: Comparison of Random Forest and Random Survival Forest Methods

**DOI:** 10.64898/2026.02.13.26346264

**Authors:** Joseph C. Vanghelof, Giorgos Tzimas, Lianlian Du, Roselyne Tchoua, Raj C. Shah

## Abstract

**Background:** When creating risk prediction models for time-to-event data, methods that incorporate time are typically used. Random survival forests (RSF), an extension of random forests (RF), are one such class of models. We compared RSF to RF in the context of time-to-event outcomes in the ASPirin in Reducing Events in the Elderly (ASPREE) randomized controlled trial. We hypothesize that RSF will have superior discrimination and calibration versus RF.

**Methods:** Participants from ASPREE residing outside the US or with missing data were excluded. A total of 2,291 participants were assigned 1:1 into training and test sets. RF and RSF models were trained using a total of 115 measures as candidate predictors. The outcome of interest was the earliest of incident dementia, physical disability, or death.

**Results:** The primary endpoint occurred in 10.5% of participants. Discrimination was similar between the models: sensitivity (∼0.75), specificity (∼0.57), positive predictive value (∼0.17), time dependent AUC (∼0.71), and Harrell’s concordance (∼0.73). Calibration was likewise similar, Brier score (∼0.09).

**Discussion:** The RF and RSF models exhibited comparable discrimination and calibration. We conclude that RSF may not always lead to more accurate predictions of outcomes compared to RF. Further examination in different clinical trial cohorts is needed to better understand the context in which adding time into outcomes risk modeling adds value.

## Background

Achieving healthy longevity is a key goal in health in the United States (US) and internationally. [1] Recent work using data from the American Community Survey conducted by the US Census Bureau highlighted an increase in the prevalence of disability-free living from 61% to 65% from 2008 to 2017 among US adults 65 and older. [2] While population-level estimates in the US are helpful, a greater benefit would come from models that can predict disability free longevity at the individual level over a fixed horizon. Such models are ideally developed using studies tracking the longitudinal trajectories of adults aged 65 and older. There is unfortunately a paucity of models that incorporate cognitive and physical function disability along with longevity, or are not based on data collected from the US, highlighting an area requiring further research and development. [3]

One such predictive model was developed using participant data from the ASPirin in Reducing Events in the Elderly (ASPREE) trial and Cox regression.[3] ASPREE was a randomized clinical trial initiated in 2010 that recruited 19,114 participants from the US and Australia and followed them over a median of 5 years.[4] It was the first large clinical trial using disability-free longevity as an outcome, measured as survival without the development of persistent physical disability or dementia. [5]

In a recent publication, a non-parametric model utilizing random forests was developed to examine heterogeneity of treatment effect, and its discrimination properties were described. [6] In that work, a discussion was raised as to whether an outcomes risk model which incorporated the element of time, rather than a random forest classifier, would have better prediction properties.

Random forest (RF) classifiers function by creating multiple decision trees, then combining the results to obtain a generally more stable prediction. [7] Each decision tree is developed using a random subset of available participants and predictors. Splits in the decision trees are determined based on whether subjects experience the event of interest or not. A modification of the RF is a random survival forest (RSF), it works similarly, but uses the log rank score as the splitting criteria to evaluate time-to-event outcomes. [8] In both RF and RSF, an ensemble prediction can be made by averaging results from all trees. [8]

Given that there is limited literature with empirical data documenting the benefit to using a model which incorporates time when working with outcomes that are time to event, we had an opportunity with the ASPREE trial to examine this issue. Here we describe in further detail the development and performance of two predictive models developed using supervised machine learning approaches. We hypothesize that a predictive model for the outcome of disability free longevity which incorporates time to event (RSF) will have superior discriminatory and calibration compared to a model trained to predict whether the outcome occurred or not (RF).

## Methods

### Source of Data

Participant level data was obtained for ASPREE trial participants. A total of 19,114 participants were recruited, 16,703 from Australia and 2,411 from the United States. The ASPREE trial was conducted in accordance with the ethical principles stated in the Declaration of Helsinki. The ASPREE-XT trial which included analyses of data from the ASPREE clinical trial was approved by The University of Iowa Institutional Review Board. Written documentation of informed consent was obtained for all participants. Clinical trial registry number: NCT01038583

### Participants

The ASPREE trial required that subjects be free of cardiovascular disease, dementia, or physical disability and at least 70 years old, or 65 if African American or Hispanic in the US. Participants were randomized to aspirin 100mg daily or placebo. The present analysis included only US participants from the ASPREE trial. Participants with any missing candidate predictors were excluded. The remaining participants were partitioned, once, equally into a training set and a test set via stratified sampling by endpoint (i.e. proportional number of endpoints in each set).

### Outcome

Our outcome of interest was disability free longevity, also called disability free survival, and was a composite of death, dementia, or physical disability, occurring prior to the end date of the trial’s intervention phase, as described in the ASPREE statistical analysis plan. [9]

### Predictors

A predictor variable set was created with 115 variables measured at baseline. Variables were selected based on having less than 5% missingness in US participants. The variables are listed in **Appendix 1**. Collinearity was limited outside of certain factors which are inherently related, such as LDL cholesterol and total cholesterol, weight and abdominal circumference, and creatinine and eGFR.

### Statistical Analysis - Model Development

Two modeling approaches were used: random forest (RF) and random survival forest (RSF). Each model was trained on the training set and 115 candidate predictors. For each approach, we developed a series of 30 random forests, with 500 decision trees per forest. To produce the ensemble risk prediction, the average result of all trees was used, with each tree having an equal vote. The forests with the median AUCs were selected as our final models. The RF and RSF models were implemented in R with the *randomForest* and *randomForestSRC* packages, respectively. [10, 11]

### Statistical Analysis - Model Assessment

The RF and RSF models were applied to subjects in the test set to compute the probability of experiencing the primary outcome at any time, and at 5 years, respectively. Overall model accuracy was measured by comparing whether the primary event was predicted (at a minimum probability of 10.5%, the mean event rate) to the observed outcome. Model discrimination ability was assessed by sensitivity, specificity, and positive predictive value (PPV) at the same risk threshold. Discrimination was further assessed using Harrell’s concordance and by computing the time-dependent area under the receiver operating characteristic curve (AUC) at 5 years after randomization. Model calibration was assessed by computing the Brier score and by grouping participants into deciles by predicted risk then plotting calibration curve of the mean predicted risk for each decile versus the observed event rate at 5 years using the Kaplan-Meier method.

### Statistical Analysis - Model Comparison

Predictions for individual subjects may be different even between models which have similar population level performance, i.e. similar discrimination and calibration. [12] Hence, four approaches were used to compare the models against each other. First, we plot for each subject the predicted risk under each model. Second, we grouped subjects in the test set into deciles by predicted risk, then compared the concordance of quantiles between the RF and RSF models. Third, we compared the variable importance for each model. Fourth, we computed the continuous absolute net classification improvement using nricens in R. [13] All other model assessment was conducted with SAS 9.4 TS1M6. [14]

## Results

### Participants

The ASPREE trial enrolled 2,411 participants from the US. After excluding 255 participants with missing data, the final study population of 2,156 was divided into a training set and a test set containing 1,078 participants each. Most participants, 89.5%, did not experience the composite endpoint, or were lost to follow-up during the study period; the median follow up time for the final study population was 5 years. Descriptive statistics for the variables which were found to have the greatest importance by the Gini index are shown in **Table 1**.

**Table 1:** Select Participant Descriptive Statistics.

|  |  | n | % |
| --- | --- | --- | --- |
| Total |  | 2156 | 100% |
| Psychoanaleptics |  | 427 | 19.8 |
| Race/Ethnicity | Black | 797 | 37.0 |
|  | Hispanic | 325 | 15.1 |
|  | Other | 48 | 2.2 |
|  | White | 986 | 45.7 |
| Psycholeptics |  | 233 | 10.8 |
| Born Overseas |  | 160 | 7.4 |
| Sex | Female | 1440 | 66.8 |
| BMI | Underweight (BMI < 18.5) | 8 | 0.4 |
|  | Normal (18.5 ≤ BMI < 25) | 504 | 23.4 |
|  | Overweight (25 ≤ BMI < 30) | 870 | 40.4 |
|  | Obese (30 ≤ BMI) | 774 | 35.9 |
| Years of Education | < 9 yrs | 103 | 4.8 |
|  | 9-11 yrs | 107 | 5.0 |
|  | 12 yrs | 441 | 20.5 |
|  | 13-15 yrs | 633 | 29.4 |
|  | 16 yrs | 393 | 18.2 |
|  | 17-21 yrs | 479 | 22.2 |
| Mineral Supplements |  | 122 | 5.7 |
| Antithrombotic Agents |  | 491 | 22.8 |
| Polypharmacy |  | 691 | 32.1 |
| Corticosteroids |  | 134 | 6.2 |
|  |  | Mean | Std Dev |
| Symbol Digit Modalities Test |  | 37.24 | 11.24 |
| Age |  | 73.89 | 5.50 |
| 3MS |  | 93.53 | 5.03 |
| SF12 Physical |  | 48.67 | 8.71 |
| Average Gait Speed (seconds per 5 meters) |  | 3.81 | 1.73 |
| Abdominal Circumference |  | 97.79 | 14.26 |
| SBP mean |  | 135.14 | 17.18 |
| eGFR CKD |  | 74.57 | 16.42 |
| Triglyceride (mmolL) |  | 1.28 | 0.64 |
| Weight |  | 79.12 | 16.94 |
| DBP Mean |  | 77.34 | 10.10 |
| SF12 Mental |  | 55.00 | 7.70 |
| Height |  | 1.65 | 0.10 |
| Creatinine |  | 81.19 | 21.80 |
| HDL (mmolL) |  | 1.57 | 0.45 |
| Average Dominant Grip Strength |  | 25.09 | 11.58 |
| HVLT Total |  | 30.49 | 7.76 |
| Total Cholesterol (mmolL) |  | 5.04 | 0.97 |
| HVLT Slope |  | 2.30 | 2.21 |

### Model Predictive Ability

Model discrimination performance is displayed in **Table 2**. Accuracy was similar between the RF (0.60) model and RSF model (0.58). Sensitivity was high in both models at 0.73 in the RF and 0.77 in the RSF model. Positive predictive value was low (0.17) in both models. The AUC was essentially equivalent (0.71) between both models, receiver operating characteristic shown in **Figure 1**. Harrell’s concordance, another measure of discrimination ability, was nearly equivalent, at 0.72 and 0.73 for the RF and RSF models, respectively. Calibration was negligibly different between the two models, with Brier scores of 0.0892 and 0.0897 for the RF and RSF models respectively. The RF model appeared well calibrated, while the RSF was biased towards predicting a higher risk than observed in low-risk deciles, and predicting a lower risk than observed in high-risk individuals, **Figure 2**.

**Table 2:** Accuracy and discriminatory performance in the test set.

| Model | Accuracy<br>(95% CI) | Sensitivity<br>(95% CI) | Specificity<br>(95% CI) | PPV<br>(95% CI) | AUC | Harrell's<br>Concordance |
| --- | --- | --- | --- | --- | --- | --- |
| Random Forest | 0.60<br>(0.57-0.62) | 0.73<br>(0.65-0.81) | 0.58<br>(0.55-0.61) | 0.17<br>(0.14-0.21) | 0.71<br>(0.66-0.76) | 0.72 |
| Random Survival<br>Forest | 0.58<br>(0.55-0.61) | 0.77<br>(0.69-0.84) | 0.56<br>(0.53-0.59) | 0.17<br>(0.14-0.20) | 0.71<br>(0.66-0.76) | 0.73 |

**Figure 1:**
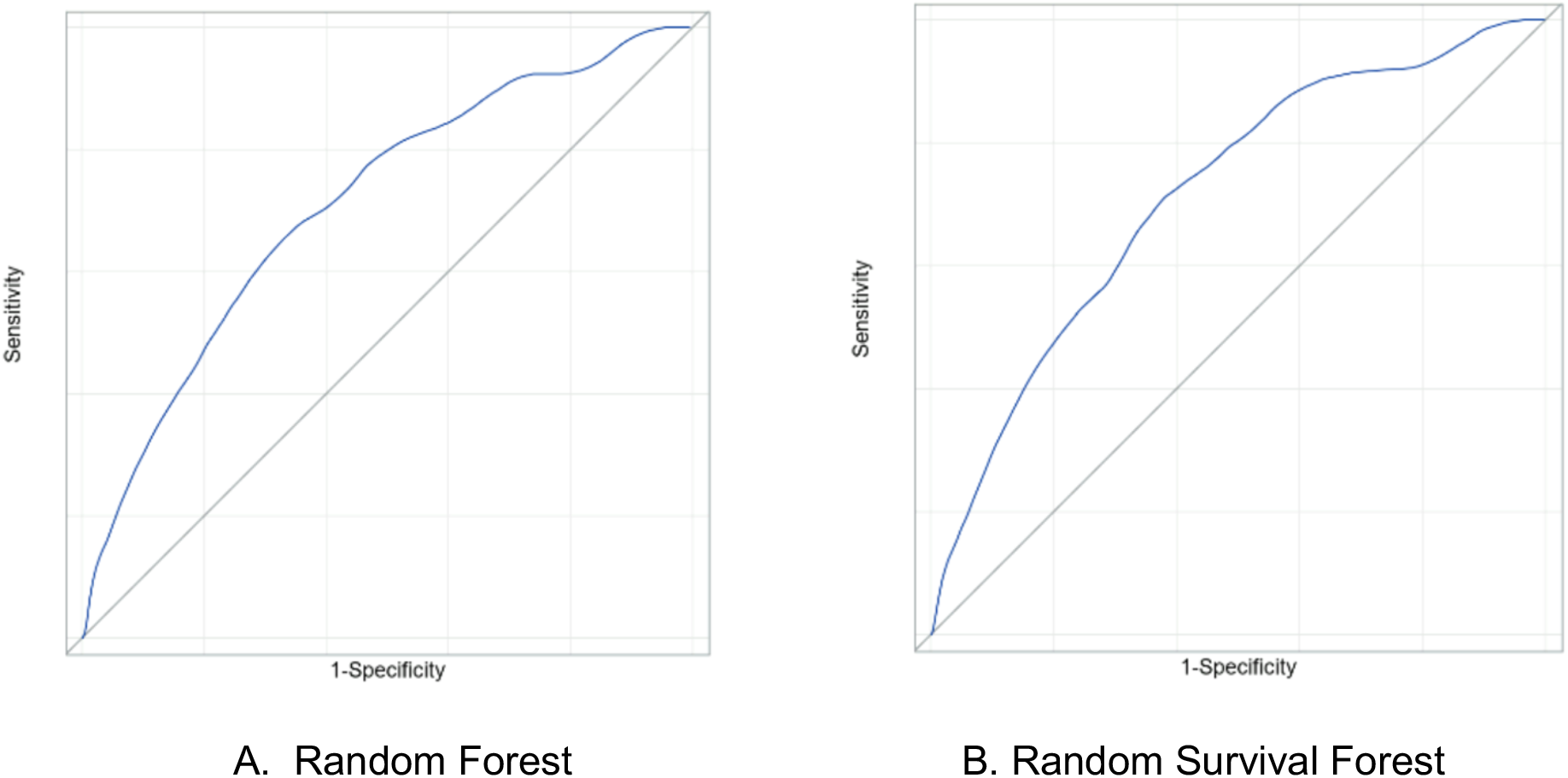
Receiver Operator Curve.

**Figure 2:**
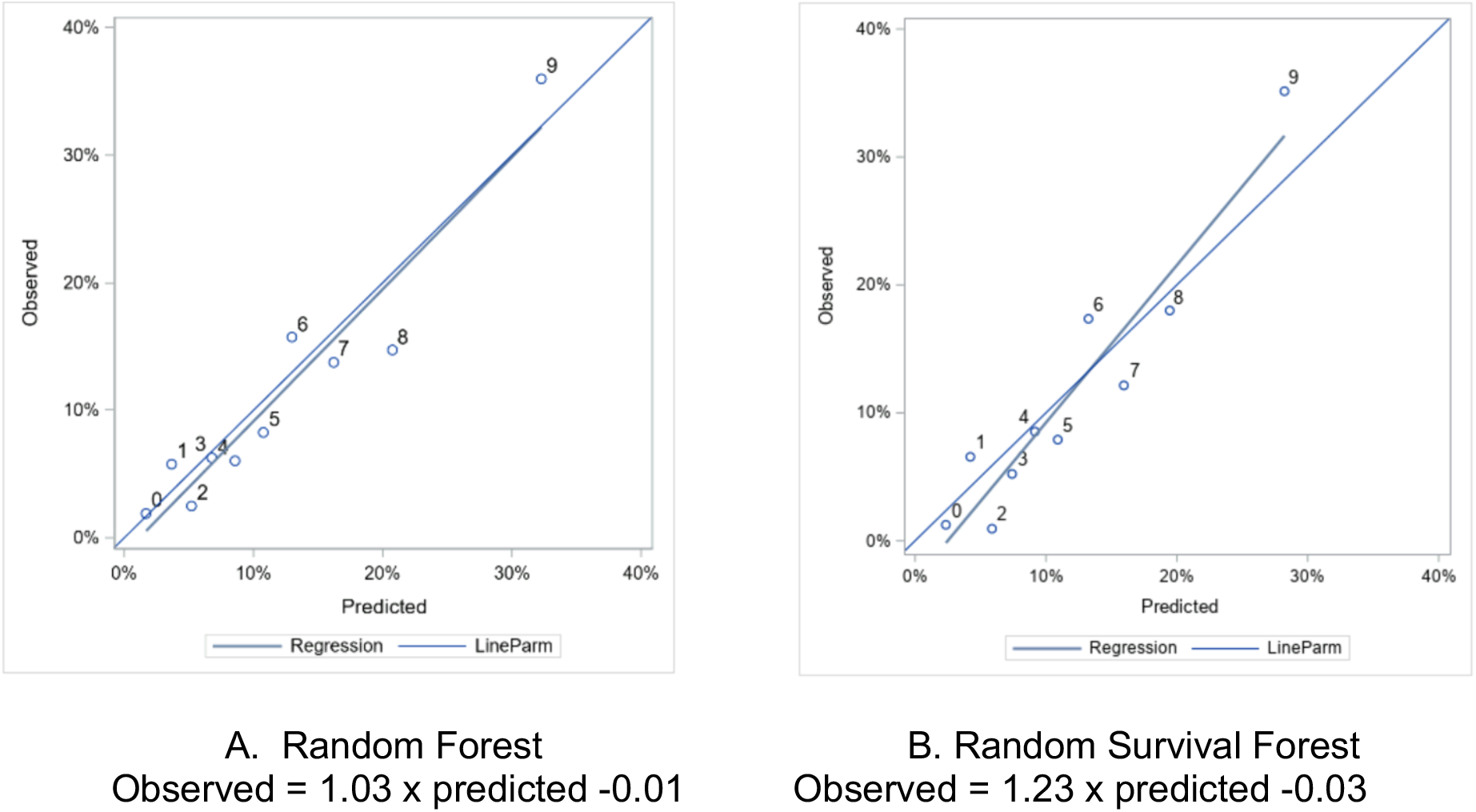
Calibration Curve, by decile of predicted risk.

### Model Comparison

At the individual level, the predicted risk for each subject was similar (R^2^ = 0.89) between the two models, **Figure 3**. When subjects in the test set were grouped into predicted risk quintiles, approximately two thirds (65%) were categorized into a concordant risk quintile, **Figure 4**. In comparing the top 30 factors for each model, 21 of the factors were represented in both models, but ranked differently, **Figure 5**. The absolute net reclassification improvement was -10.2% (95%CI -16.3% to -4.1%), Hence, 10.2% of the individuals in the study population were reclassified in the wrong direction by the RSF model compared to the RF model.

**Figure 3:**
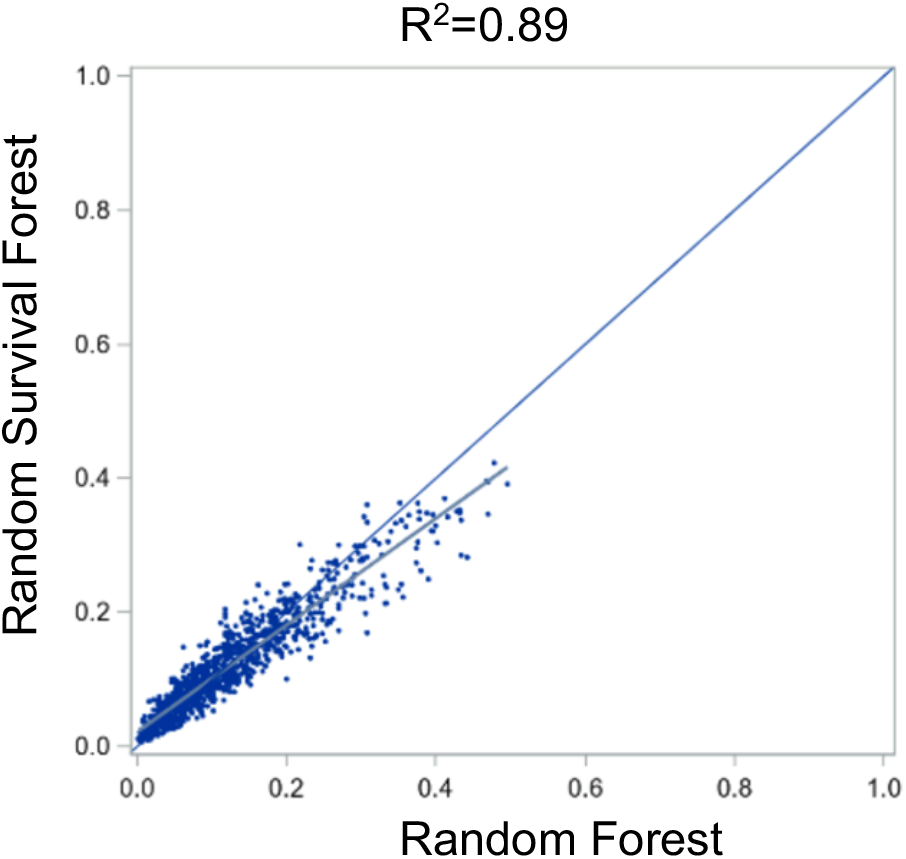
Pairwise predicted risk.

**Figure 4.**
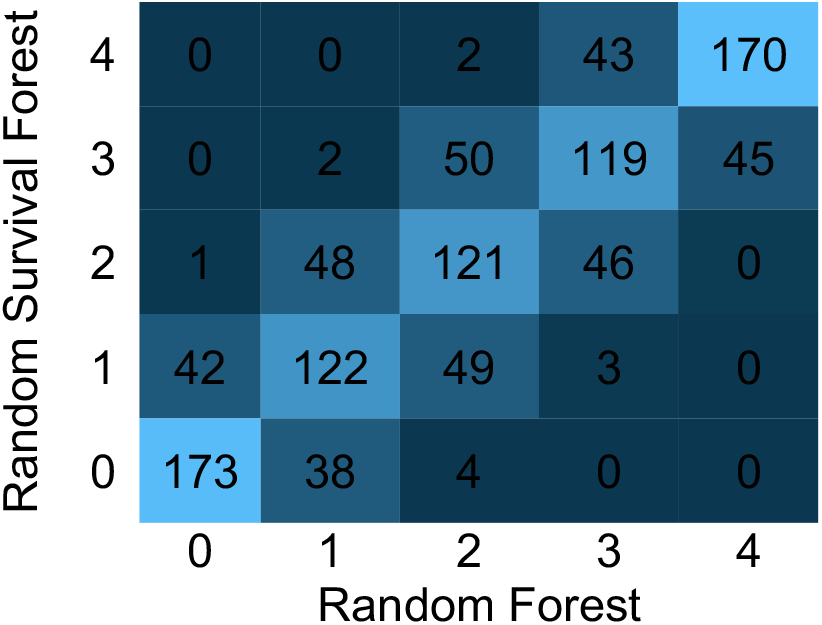
Quintile by Quintile Comparison.

**Figure 5.**
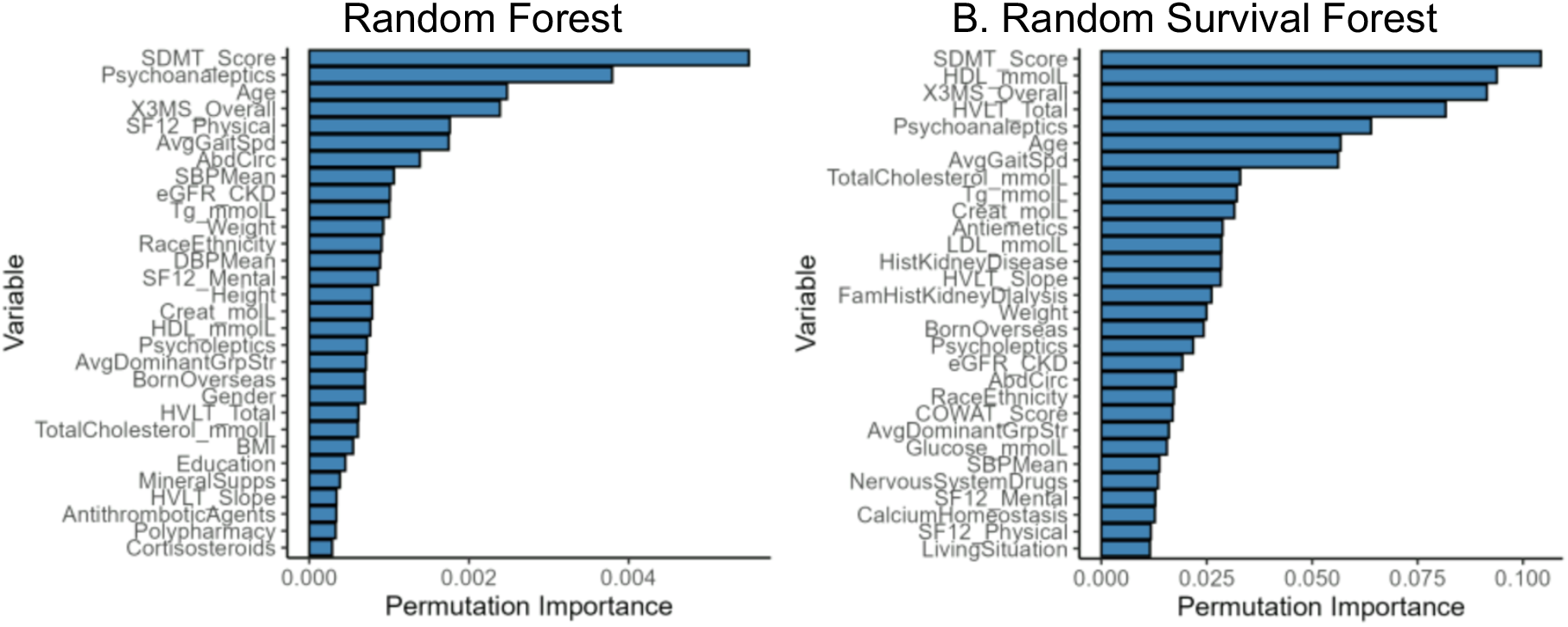
Feature importance.

## Discussion

We sought to empirically compare predictive models for time-to-event data using models that do and do not consider time. Utilizing US participant data from the ASPREE clinical trial, we compared the discrimination and calibration properties of RF and RSF models for the outcome of death, dementia, or physical disability, which occurred in 10.5% of US participants over a median of 5 years of follow-up. It is generally assumed that adding time into the models will improve outcome prediction performance for outcome risk prediction utilizing longitudinal clinical trial data. Hence, because the outcome of interest here is time-to-event, we anticipated that a model which incorporates time would produce superior predictions compared to a model that considers only whether outcome occurred or not. However, we found that the models were generally comparable in terms of discrimination and calibration. Hence, there is no evidence of improvement and possible decrease in reclassification when using the random forest model.

To put our findings in context, we searched for literature comparing models for time-to-event and binary outcomes. Odds ratios and hazard ratios each approximate risk ratios when the event is rare.[15] It is not clear whether it can be expected that a RF and a RSF will have similar predictions when the outcome is rare, though that was the case in the present analysis. It is possible the RF and RSF models performed similarly because events were rare; effectively, time-to-the-event was an unimportant aspect of the outcome and was adequately represented by whether the event occurred or not.

We conducted an additional literature search to find reviews comparing RSF to Cox regression but did not identify any. The key articles we found in this space were comparisons of individual RSF and Cox regression models, and these found that RSF outperforms Cox regression, but not in all cases. [16–20] In 2022, Neumann et al. [3] developed two Cox regression models for participants in the ASPREE trial. One model was for men, and one for women, and exhibited AUCs of 0.72 and 0.75, respectively. This is similar to the performance of the models developed in the present work with AUC ∼0.73. Notably, the models developed by Neumann et. al. were for the same outcome as our own; however, a caveat is that those models included both US and Australian participants. Hence, given the similarity of the AUCs, it seems that in this case, RF and RSF offered comparable performance to Cox regression.

Strengths of this analysis are that the data were collected as part of a randomized controlled trial and hence were carefully collected and adjudicated. Limitations of our analysis are that we did not conduct simulations to compare the modeling approaches, only one dataset was assessed, in which there was an overall low event rate, and sensitivity analyses were not performed to measure the impact of imputing missing data.

## Conclusion

Random survival forests may not always lead to more accurate predictions of outcomes compared to random forest. Other considerations need to be articulated explicitly in the decision to choose a particular model for analyses. Further work in different cohorts is needed to better understand the context in which adding time into outcomes risk modelling would add value.

## Acknowledgements

The authors recognize the significant contributions made by the research participants, staff, and investigators for the ASPirin in Reducing Events in the Elderly clinical trial.

## Authors’ contributions

JCV, GT, RT, and RCS all participated in the design of the project, critical writing of the manuscript, and data analyses. JCV, GT, RT, LD, and RCS all participated in the critical editing of the manuscript.

## Data Availability

For access to the ASPirin in Reducing Events in the Elderly (ASPREE) project data, visit ams.aspree.org. Code for this project is available at https://anonymous.4open.science/r/P428_RSF_RF.

## Declarations

### Ethics approval and consent to participate

The ASPREE-XT trial, which included analyses of data from the ASPREE clinical trial, was approved by The University of Iowa Institutional Review Board (IRB). Written documentation of informed consent was obtained for all participants. The ASPREE trial was conducted in accordance with the Declaration of Helsinki.

### Consent for publication

Not applicable

## Appendix 1: List of Candidate Predictors

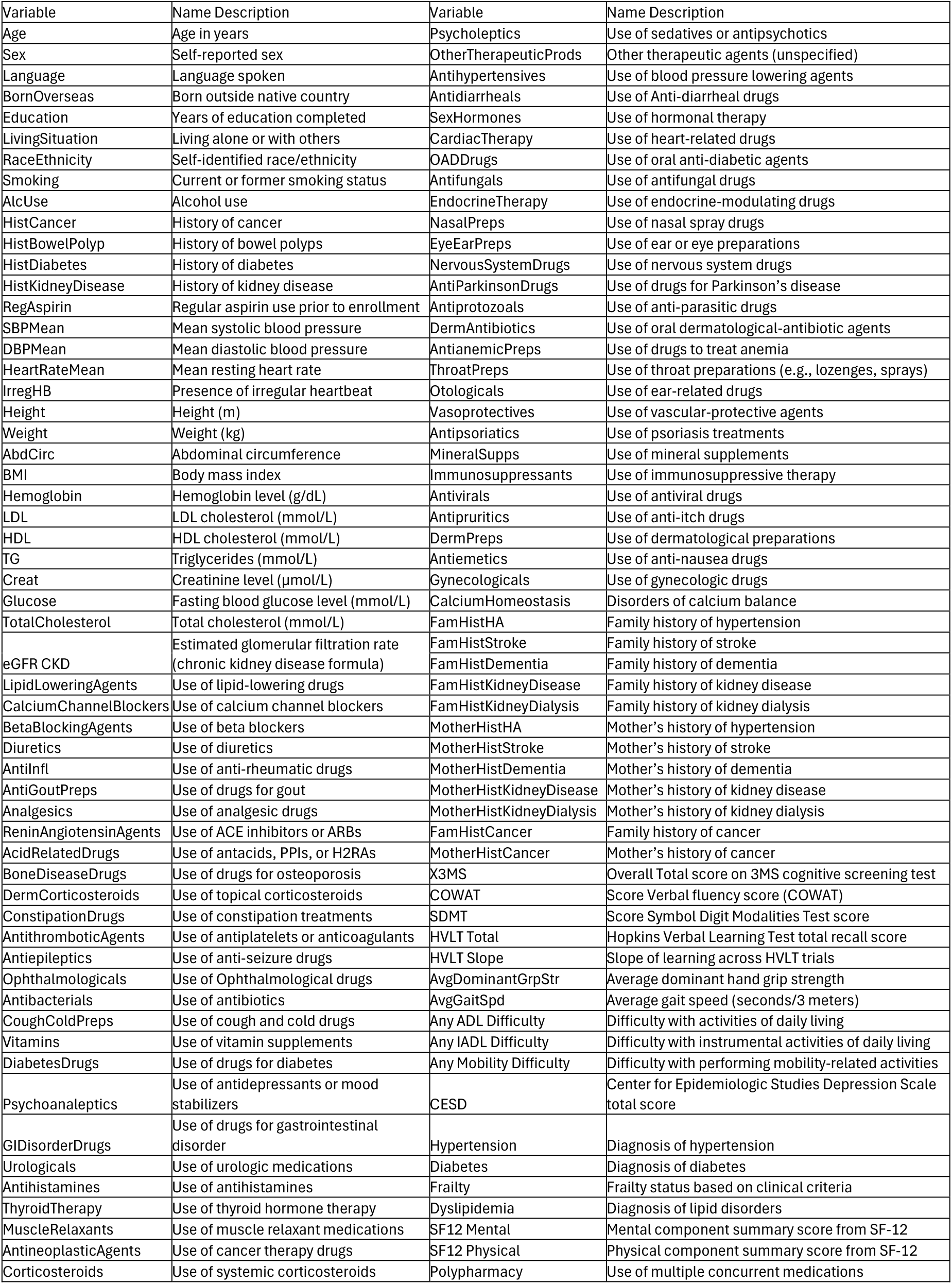

